# Loneliness among US adults in the 2024 National Health Interview Survey

**DOI:** 10.64898/2026.07.08.26357424

**Authors:** Pranta Sikder

**Affiliations:** Department of Sociology and Anthropology University of Texas at El Paso, El Paso, Texas, USA

**Keywords:** loneliness, social connection, social determinants of health, mental health, health care utilization, National Health Interview Survey, United States

## Abstract

**Rationale:** Loneliness is associated with premature mortality and poor mental health and was declared an epidemic by the US Surgeon General in 2023, yet national surveillance has relied on state-based or experimental online surveys. In 2024, the National Health Interview Survey (NHIS), the principal household health survey of the United States, measured loneliness directly for the first time.

**Objective:** To estimate the national prevalence of loneliness among US adults, identify the groups with the highest burden, and quantify associations with mental health, health status, and health care use.

**Methods:** Cross-sectional analysis of the 2024 NHIS Sample Adult file (n = 32,629; response rate 47.9%; 31,470 with valid loneliness data). Frequent loneliness was defined as feeling lonely always or usually. Survey-weighted prevalence and design-based logistic regression accounted for the complex sample design.

**Results:** In 2024, 4.9% of US adults (95% CI = 4.6, 5.2), an estimated 12.2 million people, felt lonely always or usually, and 23.7% (95% CI = 23.1, 24.3), an estimated 59.3 million, felt lonely at least sometimes. Frequent loneliness was highest among adults with family income below the federal poverty level (10.3%), adults with disability (13.6%), adults living alone (9.0%), and American Indian or Alaska Native adults (12.2%). Adults aged 65 or older had the lowest prevalence of any age group (4.0%) and adults aged 18-29 the highest (6.3%). Adjusting for sociodemographic characteristics, frequent loneliness was associated with serious psychological distress (adjusted odds ratio = 14.5; 95% CI = 12.1, 17.3), life dissatisfaction (9.0), cost-related unmet mental health care need (4.3), and emergency department use (1.8).

**Conclusions:** Loneliness among US adults is patterned by poverty, disability, and household structure rather than older age. These first NHIS estimates provide a national benchmark for monitoring loneliness and suggest that strategies for social connection should address material hardship and access to mental health care.

## 1. Introduction

Loneliness is the distressing experience that arises when a person’s social relationships are perceived to fall short of what they desire (Perlman and Peplau, 1981). It is conceptually distinct from objective social isolation and from living alone: people can be isolated without feeling lonely, and lonely while surrounded by others (Klinenberg, 2016). This subjective character makes loneliness a sensitive barometer of the adequacy, not merely the quantity, of social ties, and a growing body of evidence ties it to hypervigilance, impaired sleep, and physiological dysregulation (Hawkley and Cacioppo, 2010), to elevated risks of coronary heart disease and stroke (Valtorta et al., 2016), and to premature mortality at magnitudes comparable to established behavioral risk factors (Holt-Lunstad et al., 2015). Sociological work has long argued that social relationships should be treated as a population-level determinant of health and a target of policy in their own right (Umberson and Montez, 2010).

Policy has recently caught up with this evidence. In 2023, the US Surgeon General declared loneliness and social isolation an epidemic and issued a national strategy for social connection (Office of the U.S. Surgeon General, 2023); in 2025, the World Health Organization Commission on Social Connection identified loneliness as a leading global health concern (World Health Organization, 2025), echoing earlier warnings that loneliness is a growing public health problem worldwide (Cacioppo and Cacioppo, 2018; Surkalim et al., 2022). A National Academies consensus report likewise called for routine attention to social connection in the health care system, though its focus was confined to older adults (National Academies of Sciences, Engineering, and Medicine, 2020).

Despite this attention, US national surveillance of loneliness has been fragmented. Existing prevalence estimates come from the Behavioral Risk Factor Surveillance System, a telephone survey in which the loneliness item was fielded by a subset of states (Bruss et al., 2024), from the experimental online Household Pulse Survey, or from proprietary polls; time-use data suggest social engagement has been declining for two decades (Kannan and Veazie, 2023).

These sources differ in mode, sampling frames, and question wording, which limits comparability and leaves the country without an authoritative baseline against which to judge progress toward the goals the Surgeon General’s advisory set out.

In 2024, the National Health Interview Survey (NHIS), the principal household health survey of the US population conducted since 1957, fielded a direct loneliness question for all sampled adults for the first time (National Center for Health Statistics, 2025). The present study used those data to answer three questions. First, how prevalent is loneliness among US adults when measured in the nation’s benchmark health survey? Second, which social groups carry the greatest burden? Third, how strongly is frequent loneliness associated with mental health, health status, and health care use?

## 2. Methods

### 2.1. Study design and population

This cross-sectional study analyzed the 2024 NHIS Sample Adult public use file, released by the National Center for Health Statistics (NCHS) in June 2025 (National Center for Health Statistics, 2025). The NHIS draws a multistage probability sample of US households and interviews one randomly selected adult per household, primarily face to face with telephone follow-up. In 2024, 32,629 adults aged 18 or older completed interviews (response rate 47.9%, American Association for Public Opinion Research definition 2). The data are publicly available and de-identified; the analysis was therefore exempt from institutional review board review and informed consent requirements. The study followed the STROBE reporting guideline for cross-sectional studies.

### 2.2. Measures

Loneliness was measured with the question, “How often do you feel lonely? Would you say always, usually, sometimes, rarely, or never?” Of 32,629 respondents, 31,470 (96.4%) provided a valid response. Frequent loneliness was defined as feeling lonely always or usually, the definition used for serious loneliness in prior federal surveillance (Bruss et al., 2024); any loneliness was defined as feeling lonely at least sometimes. A companion question asked how often respondents received the social and emotional support they need.

Sociodemographic characteristics were age group, sex, race and ethnicity, educational attainment, family income as a percentage of the federal poverty level, marital status, employment in the past week, disability (a lot of difficulty or inability in at least one of six functioning domains, based on the Washington Group Short Set), household composition (living alone vs with others), urbanicity, census region, and sexual orientation. Race and ethnicity were self-reported using NHIS categories and were analyzed because the social and economic determinants of loneliness are unevenly distributed across racial and ethnic groups.

Health outcomes were serious psychological distress (Kessler 6 scale score of 13 or higher; Kessler et al., 2003); feelings of depression and of worry, nervousness, or anxiety occurring daily or weekly; dissatisfaction with life; fair or poor self-rated health; receipt of counseling or therapy from a mental health professional in the past 12 months; needing but not getting counseling or therapy because of cost in the past 12 months; and at least one emergency department visit in the past 12 months.

### 2.3. Statistical analysis

All estimates used NHIS final sample adult weights, which adjust for nonresponse and are calibrated to US population controls, so that results represent the civilian noninstitutionalized adult population (250.1 million adults). Variances were estimated by Taylor series linearization using the public use design variables for stratum and primary sampling unit, and 95% confidence intervals (CIs) for proportions used the logit transformation. Overall prevalence was also age standardized to the 2000 US standard population. Associations between frequent loneliness and each health outcome were estimated with design-based logistic regression, first unadjusted and then adjusted for age group, sex, race and ethnicity, education, family income category, and marital status. In sensitivity analyses, all models were re-estimated with any loneliness (at least sometimes) as the exposure (Appendix A). Item-level missingness was low (3.6% for the loneliness item and less than 5% for each covariate); models therefore used available cases without additional imputation, and adjusted models retained 98.4% of respondents with valid loneliness data. Statistical significance was defined as two-sided p < .05. Analyses were conducted in Python 3.10 with the samplics library (version 0.6.0).

## 3. Results

### 3.1. Prevalence of loneliness

Among the 31,470 adults with valid loneliness data (14,426 men and 17,039 women; 10,548 aged 65 or older), 4.9% (95% CI = 4.6, 5.2) felt lonely always or usually, corresponding to an estimated 12.2 million US adults, and 23.7% (95% CI = 23.1, 24.3), an estimated 59.3 million, felt lonely at least sometimes. In the full response distribution, 2.5% (95% CI = 2.3, 2.7) of adults felt lonely always, 2.4% (95% CI = 2.2, 2.6) usually, 18.8% (95% CI = 18.3, 19.4) sometimes, 25.7% (95% CI = 25.1, 26.4) rarely, and 50.6% (95% CI = 49.8, 51.4) never. The age-standardized prevalence of frequent loneliness was 5.0% (95% CI = 4.7, 5.3), similar to the crude estimate.

### 3.2. Social patterning

Frequent loneliness showed steep social gradients (Table 1; Figure 1). Prevalence among adults with family income below the federal poverty level was more than three times that of adults at or above 400% of the poverty level, and the same ratio separated adults with and without disability. Adults living alone reported more than twice the prevalence of adults living with others, and previously married and never married adults reported roughly three times the prevalence of married or cohabiting adults. Prevalence was highest among American Indian or Alaska Native adults and bisexual adults. By age, adults aged 65 or older had the lowest prevalence of any age group and adults aged 18-29 the highest. Among adults who rarely or never received needed social and emotional support, roughly one in five was frequently lonely.

**Figure 1.**
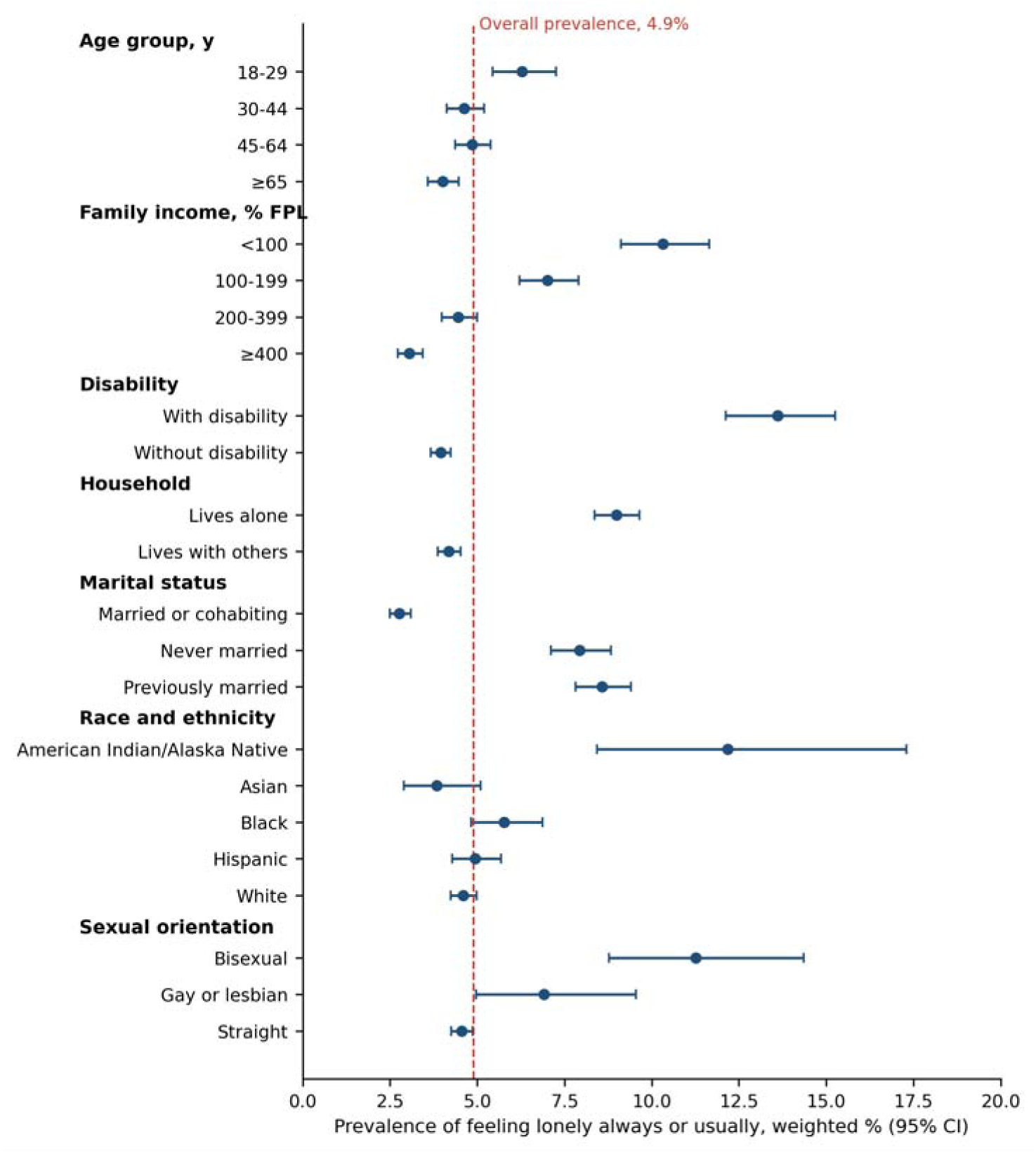
Prevalence of frequent loneliness among US adults by selected characteristics, National Health Interview Survey, 2024. Frequent loneliness defined as feeling lonely always or usually. Markers show weighted prevalence; error bars show 95% confidence intervals from Taylor series linearization; the dotted vertical line marks the overall prevalence (4.9%). Race and ethnicity categories are non-Hispanic except Hispanic; the other or multiple races category is shown in Table 1. FPL = federal poverty level. A grayscale version is provided as a separate file.

**Table 1.** Prevalence of frequent loneliness among US adults by sociodemographic characteristics, National Health Interview Survey, 2024.

| Characteristic | n (unweighted) | Lonely always or usually, weighted %<br>(95% CI) |
| --- | --- | --- |
| Overall | 31,470 | 4.9 (4.6, 5.2) |
| **Age group, years** |  |  |
| 18-29 | 3,951 | 6.3 (5.5, 7.3) |
| 30-44 | 7,374 | 4.6 (4.1, 5.2) |
| 45-64 | 9,553 | 4.9 (4.4, 5.4) |
| 65 or older | 10,548 | 4.0 (3.6, 4.5) |
| **Sex** |  |  |
| Men | 14,426 | 4.7 (4.3, 5.2) |
| Women | 17,039 | 5.1 (4.7, 5.5) |
| **Race and ethnicity (self-reported)** |  |  |
| American Indian or Alaska Native, non-Hispanic | 506 | 12.2 (8.4, 17.3) |
| Asian, non-Hispanic | 1,785 | 3.8 (2.9, 5.1) |
| Black, non-Hispanic | 3,082 | 5.8 (4.8, 6.9) |
| Hispanic | 4,561 | 4.9 (4.3, 5.7) |
| White, non-Hispanic | 21,140 | 4.6 (4.2, 5.0) |
| Other or multiple races, non-Hispanic | 396 | 7.1 (4.6, 10.7) |
| **Educational attainment** |  |  |
| Less than high school | 3,311 | 7.7 (6.7, 8.9) |
| High school diploma or GED | 7,278 | 5.3 (4.7, 6.0) |
| Some college | 8,759 | 5.1 (4.6, 5.7) |
| Bachelor's degree or higher | 11,993 | 3.2 (2.8, 3.6) |
| **Family income, % of federal poverty level** |  |  |
| <100 | 3,278 | 10.3 (9.1, 11.7) |
| 100-199 | 5,570 | 7.0 (6.2, 7.9) |
| 200-399 | 9,399 | 4.5 (4.0, 5.0) |
| 400 or higher | 13,223 | 3.1 (2.7, 3.4) |
| **Marital status** |  |  |
| Married or cohabiting | 16,442 | 2.8 (2.5, 3.1) |
| Never married | 6,736 | 7.9 (7.1, 8.8) |
| Previously married | 8,103 | 8.6 (7.8, 9.4) |
| **Employment, past week** |  |  |
| Employed | 17,236 | 4.1 (3.7, 4.5) |
| Not employed | 14,063 | 6.2 (5.7, 6.7) |
| **Disability status** |  |  |
| With disability | 3,583 | 13.6 (12.1, 15.3) |
| Without disability | 27,886 | 4.0 (3.7, 4.2) |
| **Household composition** |  |  |
| Lives alone | 10,150 | 9.0 (8.4, 9.7) |
| Lives with others | 21,320 | 4.2 (3.9, 4.5) |
| **Urbanicity** |  |  |
| Large central metropolitan | 8,988 | 4.9 (4.4, 5.4) |
| Large fringe metropolitan | 6,566 | 4.0 (3.5, 4.7) |
| Medium or small metropolitan | 8,807 | 5.2 (4.7, 5.8) |
| Nonmetropolitan | 7,109 | 5.7 (4.9, 6.6) |
| **Census region** |  |  |
| Midwest | 7,310 | 4.7 (4.1, 5.3) |
| Northeast | 4,907 | 4.4 (3.7, 5.3) |
| South | 11,401 | 5.1 (4.6, 5.6) |
| West | 7,852 | 5.1 (4.5, 5.7) |
| **Sexual orientation** |  |  |
| Bisexual | 753 | 11.3 (8.8, 14.4) |
| Gay or lesbian | 667 | 6.9 (5.0, 9.6) |
| Straight | 29,399 | 4.6 (4.3, 4.9) |
| Something else | 384 | 10.5 (7.5, 14.5) |
| **Receipt of needed social and emotional support** |  |  |
| Always, usually, or sometimes | 28,701 | 3.6 (3.3, 3.9) |
| Rarely or never | 2,683 | 19.6 (17.7, 21.6) |

Note. Frequent loneliness defined as feeling lonely always or usually. Weighted percentages with 95% confidence intervals (CIs) estimated by Taylor series linearization accounting for the complex sample design. Unweighted counts are respondents with a valid loneliness response; category totals differ from 31,470 because of item-level missing data. Race and ethnicity were self-reported using National Health Interview Survey categories. GED = General Educational Development credential.

### 3.3. Associations with health and health care use

Frequent loneliness was associated with every health outcome examined (Table 2; all p < .001). Serious psychological distress affected 30.4% of frequently lonely adults compared with 2.4% of other adults, and one quarter of frequently lonely adults were dissatisfied with life. Frequently lonely adults more often reported fair or poor health and were more likely to have received counseling or therapy, to have gone without needed counseling or therapy because of cost, and to have visited an emergency department. Associations were attenuated but remained substantial when any loneliness was the exposure (Appendix Table A1).

**Table 2.**
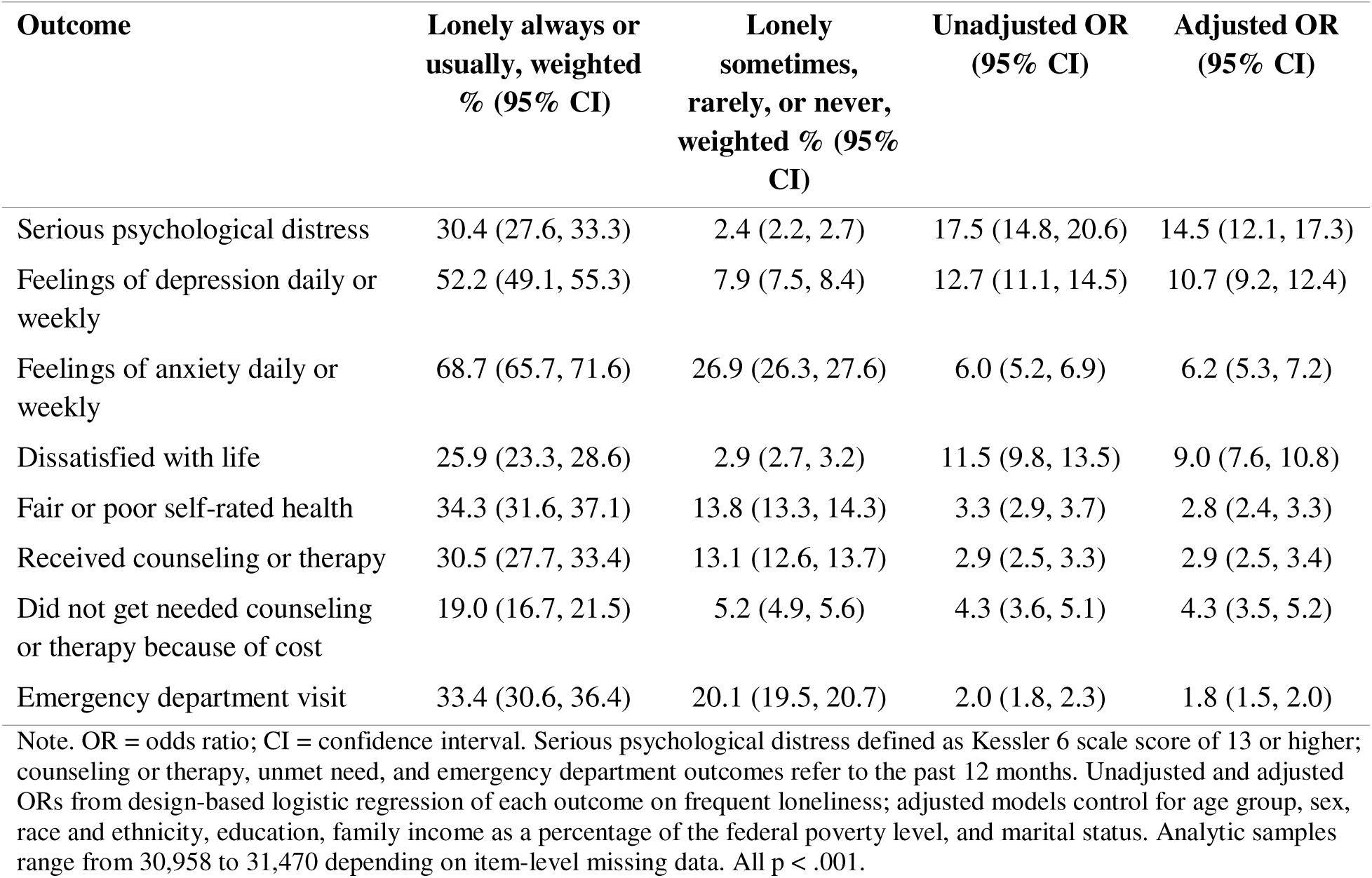
Mental health, health status, and health care use by loneliness status among US adults, National Health Interview Survey, 2024.

| Outcome | Lonely always or usually, weighted % (95% CI) | Lonely sometimes, rarely, or never, weighted % (95% CI) | Unadjusted OR (95% CI) | Adjusted OR (95% CI) |
| --- | --- | --- | --- | --- |
| Serious psychological distress | 30.4 (27.6, 33.3) | 2.4 (2.2, 2.7) | 17.5 (14.8, 20.6) | 14.5 (12.1, 17.3) |
| Feelings of depression daily or weekly | 52.2 (49.1, 55.3) | 7.9 (7.5, 8.4) | 12.7 (11.1, 14.5) | 10.7 (9.2, 12.4) |
| Feelings of anxiety daily or weekly | 68.7 (65.7, 71.6) | 26.9 (26.3, 27.6) | 6.0 (5.2, 6.9) | 6.2 (5.3, 7.2) |
| Dissatisfied with life | 25.9 (23.3, 28.6) | 2.9 (2.7, 3.2) | 11.5 (9.8, 13.5) | 9.0 (7.6, 10.8) |
| Fair or poor self-rated health | 34.3 (31.6, 37.1) | 13.8 (13.3, 14.3) | 3.3 (2.9, 3.7) | 2.8 (2.4, 3.3) |
| Received counseling or therapy | 30.5 (27.7, 33.4) | 13.1 (12.6, 13.7) | 2.9 (2.5, 3.3) | 2.9 (2.5, 3.4) |
| Did not get needed counseling or therapy because of cost | 19.0 (16.7, 21.5) | 5.2 (4.9, 5.6) | 4.3 (3.6, 5.1) | 4.3 (3.5, 5.2) |
| Emergency department visit | 33.4 (30.6, 36.4) | 20.1 (19.5, 20.7) | 2.0 (1.8, 2.3) | 1.8 (1.5, 2.0) |

Note. OR = odds ratio; CI = confidence interval. Serious psychological distress defined as Kessler 6 scale score of 13 or higher; counseling or therapy, unmet need, and emergency department outcomes refer to the past 12 months. Unadjusted and adjusted ORs from design-based logistic regression of each outcome on frequent loneliness; adjusted models control for age group, sex, race and ethnicity, education, family income as a percentage of the federal poverty level, and marital status. Analytic samples range from 30,958 to 31,470 depending on item-level missing data. All $p < .001$ .

## 4. Discussion

Using the first loneliness data ever collected in the NHIS, this study found that approximately one in 20 US adults, 12.2 million people, felt lonely always or usually in 2024, and nearly one in four felt lonely at least sometimes. To the authors’ knowledge, these are the first national loneliness estimates from the principal US household health survey; loneliness is not among the indicators in the NCHS Early Release program for 2024, and no prior peer-reviewed analysis of this measure was identified.

Two findings warrant emphasis. First, loneliness was not primarily a condition of old age. Adults aged 65 or older had the lowest prevalence of any age group and adults aged 18-29 the highest.

This pattern is consistent with life-span research showing elevated loneliness in young adulthood (Luhmann and Hawkley, 2016) and with recent US surveillance (Bruss et al., 2024), yet public discussion and many interventions remain anchored to older adults (National Academies of Sciences, Engineering, and Medicine, 2020). The results suggest that anchor is incomplete: the young adult excess observed here occurred despite young adults’ comparatively larger social networks, underscoring that loneliness reflects the perceived adequacy of relationships rather than their number (Perlman and Peplau, 1981).

Second, frequent loneliness tracked material and social disadvantage. Adults in poverty, adults with disability, and adults living alone reported prevalence two to three times the national average (see Table 1). The disability gradient mirrors findings from the United Kingdom, where the loneliness gap between adults with and without disability was widest among those who were younger, unemployed, and living in deprived circumstances (Emerson et al., 2021). These patterns support a structural reading of the loneliness epidemic: maintaining satisfying relationships requires money, health, accessible environments, and time, resources that are unequally distributed (Umberson and Montez, 2010). A national strategy for social connection that does not engage economic and disability policy is therefore likely to miss the populations where loneliness concentrates. At the same time, living alone and loneliness should not be conflated (Klinenberg, 2016): most adults living alone in these data were not frequently lonely, even though their risk was elevated.

The associations with health care use identify practical intervention points. Frequently lonely adults were substantially more likely to report cost-related unmet mental health care need and emergency department use (see Table 2). Emergency departments and safety net clinics therefore see a disproportionate share of the loneliest Americans, making them plausible settings for identifying loneliness and connecting patients to care and community resources. Because the loneliness question is scheduled to reappear in future NHIS rounds, the 2024 estimates reported here can serve as the baseline for evaluating national progress.

### 4.1. Limitations

This study has limitations. First, the design was cross-sectional, so causality cannot be inferred, and reverse causation is plausible because poor mental health can produce loneliness; the associations in Table 2 should be read as descriptive of co-occurrence, not as causal effects.

Second, loneliness was self-reported with a single direct item; direct items are widely used in surveillance and correlate with multi-item scales, but the word “lonely” carries stigma and may lead some groups, including men and older adults, to underreport, which would bias group comparisons in ways that cannot be ruled out here. Third, the response rate was 47.9%; NHIS weights adjust for nonresponse and are calibrated to population totals, but residual nonresponse bias cannot be excluded. Fourth, the survey excludes institutionalized adults, including nursing home residents, and the broad 65-or-older category may mask an increase at the oldest ages, where loneliness has been shown to rise as income, health, and partnership decline (Luhmann and Hawkley, 2016). Fifth, estimates for smaller groups, such as American Indian or Alaska Native adults, have wide confidence intervals and should be interpreted accordingly.

### 4.2. Conclusions

In 2024, an estimated 12.2 million US adults felt lonely always or usually, and 59.3 million felt lonely at least sometimes. The burden was concentrated among adults with low income, disability, or no partner and among young adults rather than older adults. These first NHIS estimates provide a national benchmark for monitoring loneliness and for evaluating the policies that the loneliness epidemic has inspired.

## Data Availability

The 2024 National Health Interview Survey public use data are freely available from the National Center for Health Statistics. Analysis code is available from the author on request.

https://ftp.cdc.gov/pub/Health_Statistics/NCHS/Datasets/NHIS/2024/adult24csv.zip

## Ethics

The study analyzed publicly available, de-identified data and was exempt from institutional review board review.

## CRediT author statement

Pranta Sikder: Conceptualization, Methodology, Formal analysis, Data curation, Writing - original draft, Writing - review and editing, Visualization.

## Acknowledgments

None.

## Appendix A. Sensitivity analysis

Appendix Table A1 re-estimates the models in Table 2 with any loneliness (feeling lonely at least sometimes; 23.7% of adults) rather than frequent loneliness as the exposure. Associations are attenuated relative to Table 2, consistent with a dose-response relationship between loneliness frequency and health, but remain substantial and statistically significant (all p < .001) for every outcome.

**Appendix Table A1.** Associations between any loneliness (at least sometimes) and health outcomes, National Health Interview Survey, 2024.

| Outcome | Unadjusted OR<br>(95% CI) | Adjusted OR (95%<br>CI) |
| --- | --- | --- |
| Serious psychological distress | 13.5 (11.3, 16.2) | 11.8 (9.7, 14.3) |
| Feelings of depression daily or weekly | 8.6 (7.7, 9.5) | 7.8 (7.0, 8.7) |
| Feelings of anxiety daily or weekly | 4.3 (4.0, 4.6) | 4.6 (4.2, 4.9) |
| Dissatisfied with life | 6.4 (5.5, 7.4) | 5.4 (4.6, 6.4) |
| Fair or poor self-rated health | 2.4 (2.2, 2.6) | 2.3 (2.1, 2.5) |
| Received counseling or therapy | 3.2 (2.9, 3.4) | 3.1 (2.9, 3.4) |
| Did not get needed counseling or therapy<br>because of cost | 4.6 (4.0, 5.2) | 4.5 (3.9, 5.2) |
| Emergency department visit | 1.5 (1.4, 1.7) | 1.4 (1.3, 1.5) |

Note. OR = odds ratio; CI = confidence interval. Unadjusted and adjusted ORs from design-based logistic regression; adjusted models control for age group, sex, race and ethnicity, education, family income as a percentage of the federal poverty level, and marital status. Analytic samples range from 30,958 to 31,470. All $p < .001$ .

